# Randomised evaluation platform – interventions to treat older people with sarcopenia (REVITALiSE): protocol and description of intervention selection process

**DOI:** 10.1101/2025.07.04.25330887

**Authors:** Claire McDonald, Philippa Watts, Helen Atkinson, Katie Kucukcan, Aritra Mukherjee, Mosammat Polyma, Avan A Sayer, Gemma F Spiers, Sarah Khan, Jane Nesworthy, Emily G Robertson, Hannah O’Keefe, James M S Wason, Nina Wilson, Miles D Witham

## Abstract

**Introduction:** Sarcopenia is the age-related loss of muscle mass and strength. It is associated with significant adverse personal and health-economic outcomes. Despite advances in understanding the biology of muscle ageing, effective treatments remain limited. Exercise is currently the only proven intervention, but many older people are unable or unwilling to sustain the intensity of exercise required to gain results. Consequently, there is a major unmet need for new therapies. REVITALiSE is an early-phase experimental medicine platform trial designed to efficiently evaluate promising interventions in people with sarcopenia. By identifying and selecting the most promising interventions to progress to large randomised controlled trials, REVITALiSE aims to accelerate the development of effective therapies for this under-served population.

**Methods and analysis:** The REVITALiSE platform comprises a series of parallel-group, individually randomised, controlled, open-label, proof-of-concept subtrials. Each subtrial will enrol 30 participants aged 65 years and older with probable sarcopenia, defined according to the European Working Group on Sarcopenia in Older People (EWGSOP) guidelines. The platform is designed to evaluate a range of interventions, including exercise-based approaches, device-based therapies, and nutraceuticals. Participants will be randomised in a 1:1 ratio to receive either the intervention or usual care. The primary outcome, analysed in a modified intention to treat (mITT) population, is the between-group difference in four-metre walking speed between baseline and 12-week follow-up. Secondary outcome measures specified in the master protocol include handgrip strength, the Short Physical Performance Battery (SPPB), and lean muscle mass (assessed by Dual X-ray absorptiometry). Muscle biopsies of the vastus lateralis will also be taken at baseline and follow-up. Additional mechanistic outcomes will be determined by the proposed mode of action of each intervention and specified in the relevant subtrial annex. Adverse events will be recorded for the duration of the trial.

**Ethics and dissemination:** UK Health Research Authority and Northeast – Tyne and Wear South Research Ethics Committee (IRAS 352708). Results will be made available to participants, their families, patients with sarcopenia, the public, regional and national clinical teams, and the international scientific community.

Trial registration number: ISRCTN10801475

Protocol V1.0 08 May 2025

Sponsor: Newcastle Upon Tyne NHS Foundation Trust

REC: NE Tyne and Wear South NHS Research Ethics Committee ref: (IRAS 352708; ref: 25/NE/0115)

## Introduction and background

Sarcopenia is the progressive age-related loss of skeletal muscle strength, mass and function (1). It affects 10-27% of people aged over 60 (2) and is strongly associated with a number of adverse clinical outcomes, including increased risk of falls, functional decline, hospitalisation, loss of independence, and premature mortality. Sarcopenia also leads to a substantial socioeconomic burden driven by greater healthcare utilisation, prolonged hospitalisation and rehabilitation, and escalating long-term care requirements(3), with direct costs to the UK National Health Service estimated at £2.5 billion per year.(4)

The pathophysiology of sarcopenia is multifactorial and not fully understood, but rapid progress in the field of skeletal muscle biology has identified several contributory and interlinked biological processes (5). These include mitochondrial dysfunction, oxidative stress, cellular senescence, autophagy, altered nutrient sensing, chronic inflammation and hormonal changes (e.g. reduced levels of testosterone). These factors interact to contribute to muscle protein breakdown, decreased muscle synthesis and loss of muscle mass. Additionally, denervation plays a critical role, with progressive loss of motor neurons and alterations to the neuromuscular junction, which impair muscle contractility and contribute to muscle atrophy.

Despite a growing understanding of the biological mechanisms underlying sarcopenia, the therapeutic options available remain limited. Resistance exercise remains the only intervention with consistent, high-quality evidence supporting its efficacy in slowing or reversing the progression of sarcopenia [Hurst 2022]. However, many older individuals are either unable or unwilling to engage in the intensity or duration of exercise required to achieve therapeutic benefit. Even where resistance training is undertaken, it may only be partially successful in reversing the deleterious effects of sarcopenia, and thus finding combination therapies that augment the effect of exercise is also desirable. New therapies (biochemical or device-based approaches) that can augment or substitute for exercise-based regimens are therefore needed (3).

The translation of basic research in sarcopenia into clinical therapies has been slow and largely disappointing (6). This translational gap in arises from a combination of issues related to preclinical research models(3), a lack of a standardised or systematic approach for identifying and prioritising preclinical interventions that are most likely to succeed in clinical trials, traditional clinical trial designs that are not suited to people with sarcopenia(7) and diagnostic barriers that result in recruitment challenges (8, 9). While numerous biological targets and potential therapies have been identified in preclinical models, there is often no clear framework for selecting which interventions to advance.(10) This is compounded by limitations of our current animal models of sarcopenia, which are predominantly rodent-based, lack a standardised approach, and often fail to replicate the full complexity of human sarcopenia. These models may not accurately reflect the multifactorial nature of the condition in humans, including the presence of comorbidities and polypharmacy (3, 11).

Clinical trials in sarcopenia are inherently challenging. Sarcopenia remains under-diagnosed in clinical practice, and even when it is recognised, it is often not coded in electronic health records (EHRs), complicating the identification of eligible participants for clinical studies (12). Furthermore, sarcopenia is not yet widely recognised by the public. Many older adults with sarcopenia are unaware of their condition, and even if diagnosed, they may not fully comprehend its implications or the necessity for treatment.(12) This lack of awareness can reduce the likelihood of participating in clinical research. Moreover, a significant proportion of these individuals live with multimorbidity and polypharmacy (3, 13). Stringent eligibility criteria intended to safeguard patient safety often result in underrepresentation of these groups in clinical trials (14, 15). Most clinical trials for sarcopenia use traditional designs that may not be ideal for this complex, multifactorial condition. Recruitment is often slow due to challenges identifying participants, and retention has been suboptimal, likely reflecting the fact that most sarcopenia trials are not designed to accommodate the challenges faced by patients with sarcopenia, such as mobility difficulties, comorbidities, and polypharmacy. Innovation is particularly needed in early-phase trials, where novel interventions are tested for proof-of-concept in humans in order to make the case for phase II (efficacy) and phase III (effectiveness) trials. Without a thriving pipeline of proof-of-concept studies, later-phase trials cannot be justified or funded.

Platform trials offer a highly efficient approach to evaluating multiple therapeutic interventions simultaneously under a single protocol. (16) This is particularly advantageous in sarcopenia, a multifactorial condition with diverse biological targets and a wide range of candidate treatments— including pharmacological agents, nutraceuticals, and exercise-based interventions. Unlike traditional trial designs, platform trials allow interventions to be added, modified, or discontinued without setting up an entirely new trial with the attendant long lead-times and duplication of effort that each new trial brings. The inclusion of multiple interventions can provide a broader choice for potential participants. Such approaches can be more cost-effective, leveraging shared infrastructure and resources to reduce duplication and overheads (17).

In this paper, we present the master protocol for the REVITALiSE sarcopenia platform trial – a novel, early-phase sarcopenia platform trial designed to evaluate a range of non-pharmacological interventions for older people with sarcopenia. We also present a process for identifying and selecting potential interventions for testing in early-phase sarcopenia trials, which we have used to prioritise interventions for evaluation in REVITALiSE.

## Methods

### Trial design, aims and objectives

The REVITALiSE sarcopenia platform trial consists of multiple parallel group, randomised controlled subtrials. Participants with muscle weakness, operationalised as low grip strength or prolonged five-times sit-to-stand time will be allocated to an active subtrial on the platform and then randomised to either the intervention or usual care. Treatments will be open-label with blinded outcome assessment. The overall aim of the REVITALiSE sarcopenia platform trial is to provide proof of concept evidence that different interventions may be able to improve physical performance in people with sarcopenia. There are three aspects to the generation of this proof-of-concept evidence:

A) To ascertain whether the intervention can improve target biological pathways relevant to sarcopenia (signal of mechanistic effect)
B) To provide indicative data that the intervention may be able to improve physical performance in people with sarcopenia (signal of efficacy)
C) To provide initial data that the intervention can be delivered safely to people with sarcopenia and that the intervention is tolerated (signal of feasibility/tolerability)

### Trial setting

Participants will be recruited from the North-East of England. The trial is hosted by the Newcastle upon Tyne Hospitals NHS Foundation Trust, with additional participant identification centres (PICs) in primary and secondary care organisations in the North-East of England region. In addition, participants will be sought from existing registries (the SarcNet sarcopenia registry(18), the MULTIPLE registry of participants with multiple long-term conditions, and from the NIHR BioResource (19) Follow-up will be for a total of 12 weeks, with outcomes measured at baseline and 12 weeks. Safety follow-up visits will be conducted as specified in the subtrial annexes between the baseline and follow-up visits. Randomisation in each subtrial will be performed via a central, web-based allocation system (Research Electronic Data Capture; REDCap [https://projectredcap.org]) hosted at Newcastle Upon Tyne NHS Hospitals Trust.

### Eligibility criteria

The target population for REVITALiSE is older people with probable sarcopenia – a status operationalised in the second European Working Group on Sarcopenia in Older People (EWGSOP2) guidelines as the presence of low muscle strength with or without the presence of low muscle mass. Use of this diagnostic definition and the EWGSOP2 diagnostic thresholds will align the trial population with the way that sarcopenia is currently diagnosed in clinical practice in Europe and has been successfully used in previous sarcopenia trials (20, 21). The key inclusion and exclusion criteria for the platform master protocol are outlined in Figure 1. These criteria are designed to ensure the inclusion of participants with sarcopenia, while excluding those with other underlying causes of muscle weakness that could confound the interpretation of outcomes. They also take into account factors relevant to the feasibility and safety of obtaining muscle biopsies, ensuring that enrolled participants are suitable for the procedures required within the platform. Individual subtrials conducted within the platform may include additional inclusion or exclusion criteria specific to the intervention being evaluated. These criteria are designed to ensure participant safety and the appropriateness of the intervention for the target population. All subtrial–specific criteria will be detailed in the relevant subtrial annexes accompanying the master protocol.

**Figure 1.**
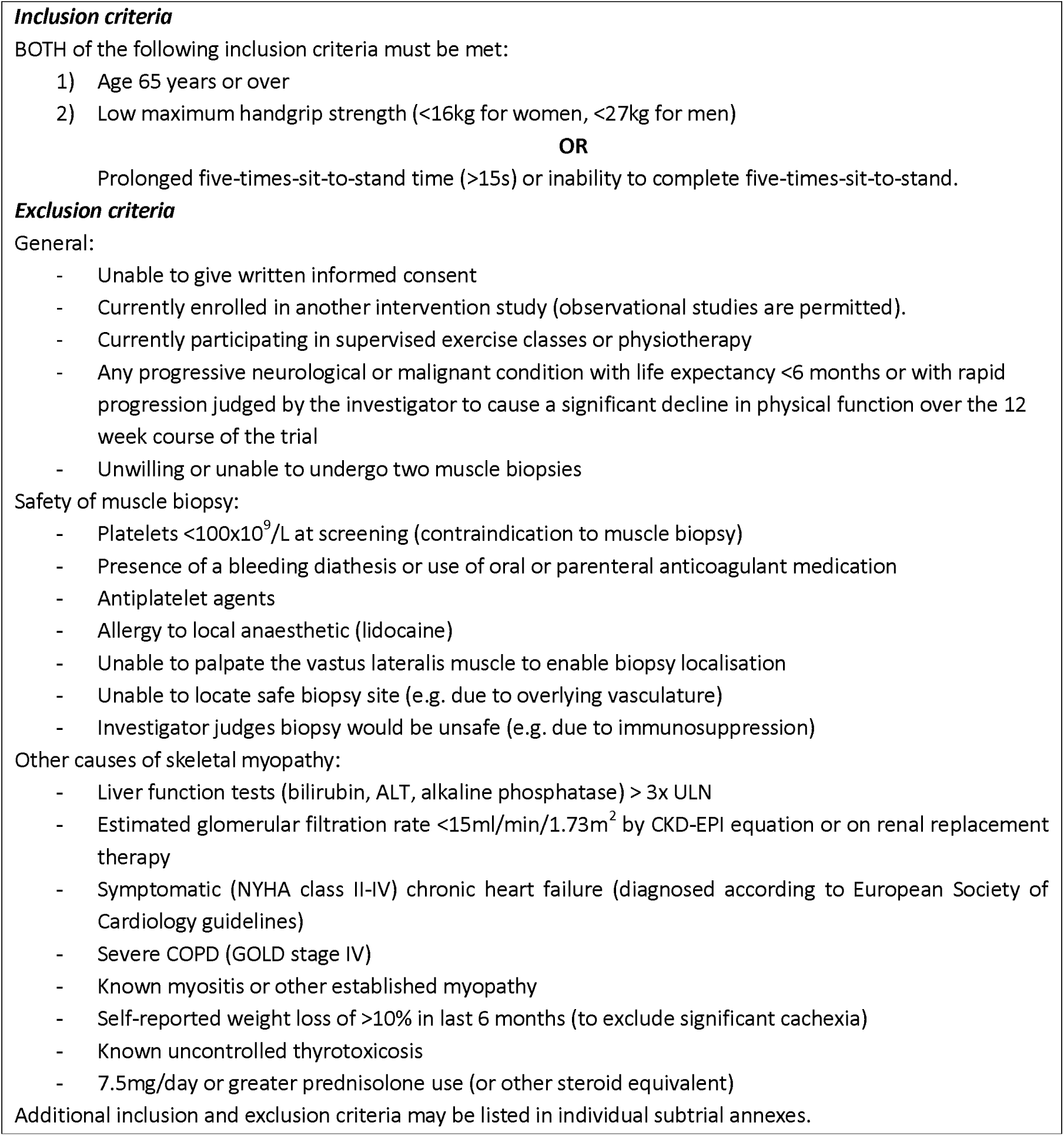
REVITALiSE master protocol inclusion and exclusion criteria:

**Figure 2.**
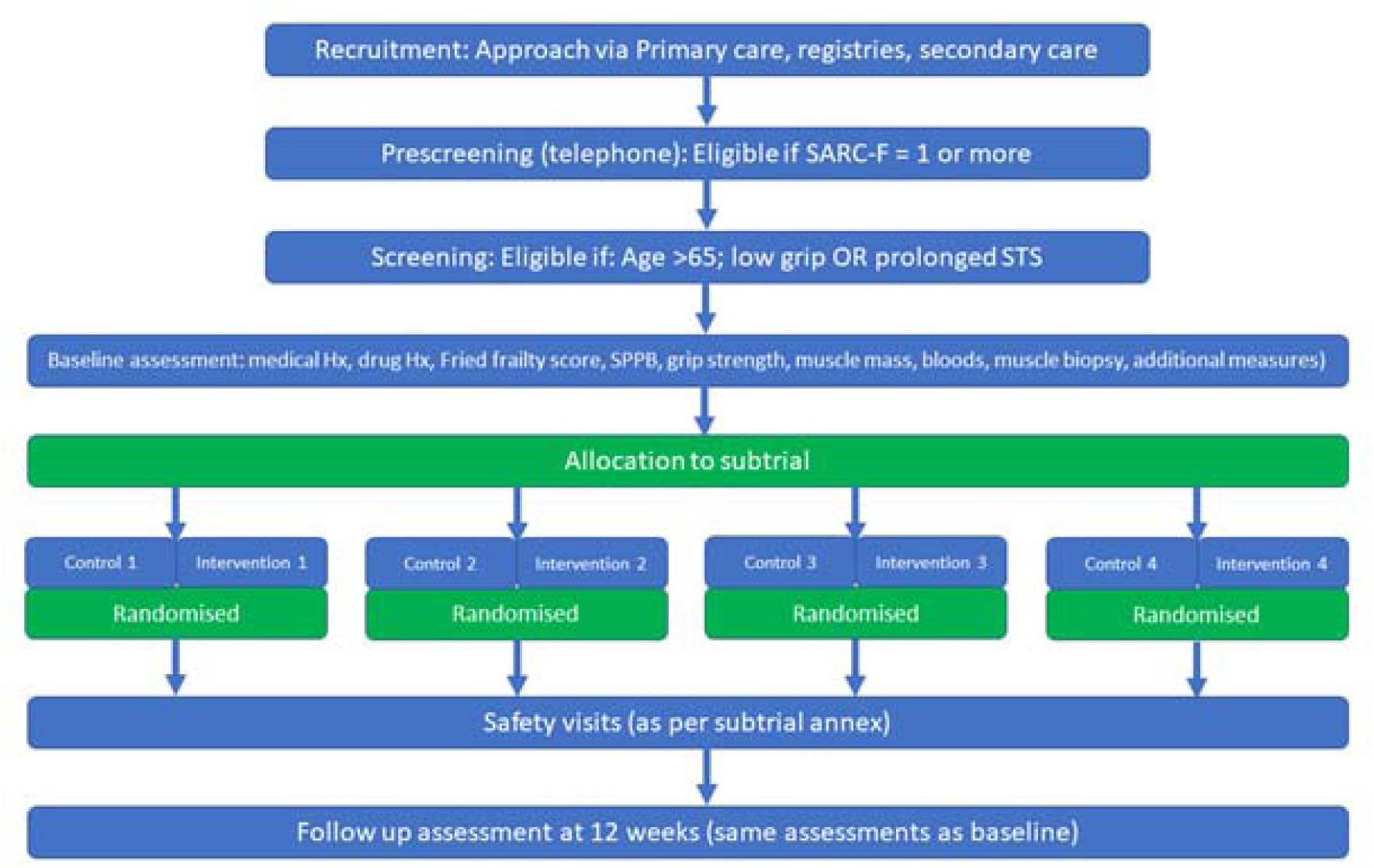
Flow diagram for the REVITALiSE platform trial.

**Figure 3.**
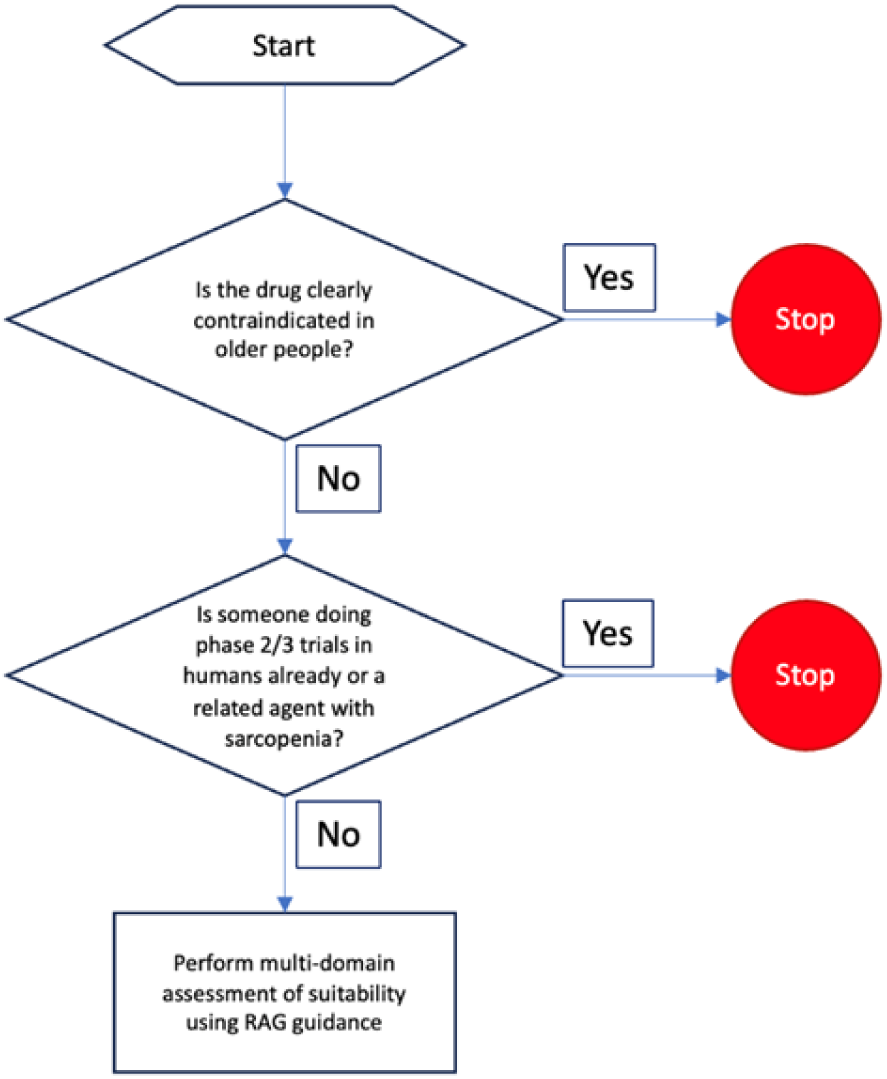
Intervention selection flowchart.

### Interventions

REVITALiSE is designed to evaluate a broad range of non-pharmacological interventions. It is not designed to study drug interventions and thus does not have approval as a Controlled Trial of Investigational Medicinal Products (CTIMP). It has the capacity to evaluate nutraceuticals, whole-food interventions, exercise and selected device interventions. The use of separate subtrials rather than a single control group with multiple intervention arms provides the flexibility to test very different types of interventions whilst still enabling the logistical efficiencies of the platform design to be realised. It also enables pooling of control groups where appropriate. Further information on the intervention selection process for REVITALiSE is given in the section below.

### Outcomes

#### Primary outcome

The primary outcome for each REVITALiSE subtrial is the between-group difference in 4-metre walk speed at 12 weeks, adjusted for baseline values. Amendments to the primary outcome are permitted as part of each subtrial annex, but the outcomes must be specified prior to the commencement of each subtrial. The 4 metre walk speed is a well-studied marker of integrated whole-body physiology, which strongly predicts future dependency, falls and death(22, 23). It is a marker of the severity of sarcopenia recommended by the EWGSOP2 guidelines. Furthermore, it is a stable and reproducible measure but is also responsive to change with interventions, including in those with sarcopenia(24).

#### Secondary outcomes

Secondary outcomes are categorised into three domains: indicators of efficacy, measures of safety and tolerability, and assessments of underlying biological mechanisms

- Signals of Efficacy To assess preliminary signals of efficacy, between-group differences at 12 weeks will be evaluated using maximal handgrip strength, the Short Physical Performance Battery (SPPB), and lean body mass. In addition, seven-day remote monitoring with a wearable device (AX6; Axivity Ltd, Newcastle upon Tyne, UK) will be employed to capture detailed measures of walking activity, gait speed, gait variability, and postural control at both baseline and follow-up.
- Safety and tolerability outcomes Safety will be assessed by analysing adverse events among all participants who receive at least one dose of the intervention. Adverse events will be recorded and categorised according to severity and relatedness to the intervention, with safety data reviewed regularly by the trial management team.
- Mechanistic Outcomes Mechanistic outcomes will be selected based on the hypothesised mode of action of the intervention and defined within the relevant subtrial annex. In most cases, these will include measures such as muscle fibre cross-sectional area and the distribution of muscle fibre types (Type I, IIa, and IIx), alongside biomarkers relevant to biological ageing and sarcopenia. For example, if the intervention is thought to exert senolytic effects, mechanistic endpoints will include circulating markers of the senescence-associated secretory phenotype (SASP) and skeletal muscle transcriptomic signatures associated with cellular senescence. Conversely, for interventions targeting mitochondrial function, mechanistic outcomes may include measures such as mitochondrial energy metabolism (e.g., via ³¹P magnetic resonance spectroscopy or near-infrared spectroscopy [NIRS]), mitochondrial DNA copy number, and enzymatic activity of mitochondrial complexes I–IV. Full details of the mechanistic outcomes for each subtrial will be published in the corresponding annex prior to trial initiation.

### Participant timeline

#### Pre-screening

Participants expressing interest in the trial will undergo a brief pre-screening process to assess provisional eligibility, which will be conducted by telephone or in person. Verbal consent will be obtained for the pre-screen process, which includes access to medical records for further review of suitability for the trial. Participants will be asked if they have any diagnosis listed as exclusion criteria (whether for the overall platform or any current subtrial), and what medications they are on. They will also be asked if they are willing to undergo two muscle biopsies. The SARC-F questionnaire will be administered, comprising five questions on physical function, with a score ranging from 0 to 10. A score of 1 or more will be used to denote an elevated probability of having sarcopenia and enable progression to the screening visit. Participants who are eligible at pre-screening will be given or posted the full participant information sheet (PIS) and will have at least 48 hours to consider their participation.

#### Screening

Informed, written consent for participation in the platform trial will be obtained at the screening visit, prior to conducting any trial-related procedures, including screening assessments. Although consent for a specific subtrial will not be obtained until after the screening visit is completed, the visit will include a discussion of the currently active subtrials. This will allow participants to make an informed decision about trial entry based on their likely eligibility and preferences regarding available interventions. Once initial consent is given, demographic data, medical history, and current medication use will be recorded. Screening assessments will include measurement of maximal handgrip strength and five-times sit-to-stand time, as well as inspection and palpation of the vastus lateralis muscle to assess suitability for biopsy. Blood samples will be collected for full blood count (FBC), urea and electrolytes (U&Es), and liver function tests (LFTs). Participants will proceed to the baseline visit and randomisation only if they are confirmed as eligible following these assessments. All participants must meet the core eligibility criteria for the main platform trial and must also meet the eligibility criteria for at least one active subtrial. Ineligibility for one or more subtrials does not preclude participation in others, provided the platform criteria and at least one subtrial’s criteria are fulfilled. Once subtrial eligibility is confirmed, additional informed consent specific to the selected subtrial will be obtained before randomisation

#### Study Visits

The schedule of events is given in Table 1. Main trial outcomes will be assessed across two baseline visits and two follow-up visits, each set scheduled seven days apart to accommodate accelerometer data collection. The first baseline visit includes key assessments and accelerometer fitting, followed by a second baseline visit one week later, during which the muscle biopsy is performed and the accelerometer is collected. Similarly, follow-up outcomes will be assessed during two final visits: the first at 12 weeks post-randomisation and a second visit seven days later. A safety visit will be conducted four weeks after the initial baseline visit and may include repeat blood tests and review of any adverse events. Participants will also receive follow-up phone calls two and seven days after the second baseline and follow-up visits to assess their recovery following the muscle biopsy. A final follow-up phone call will be conducted four weeks after trial completion to monitor for any delayed or late-emerging adverse events

**Table 1.**
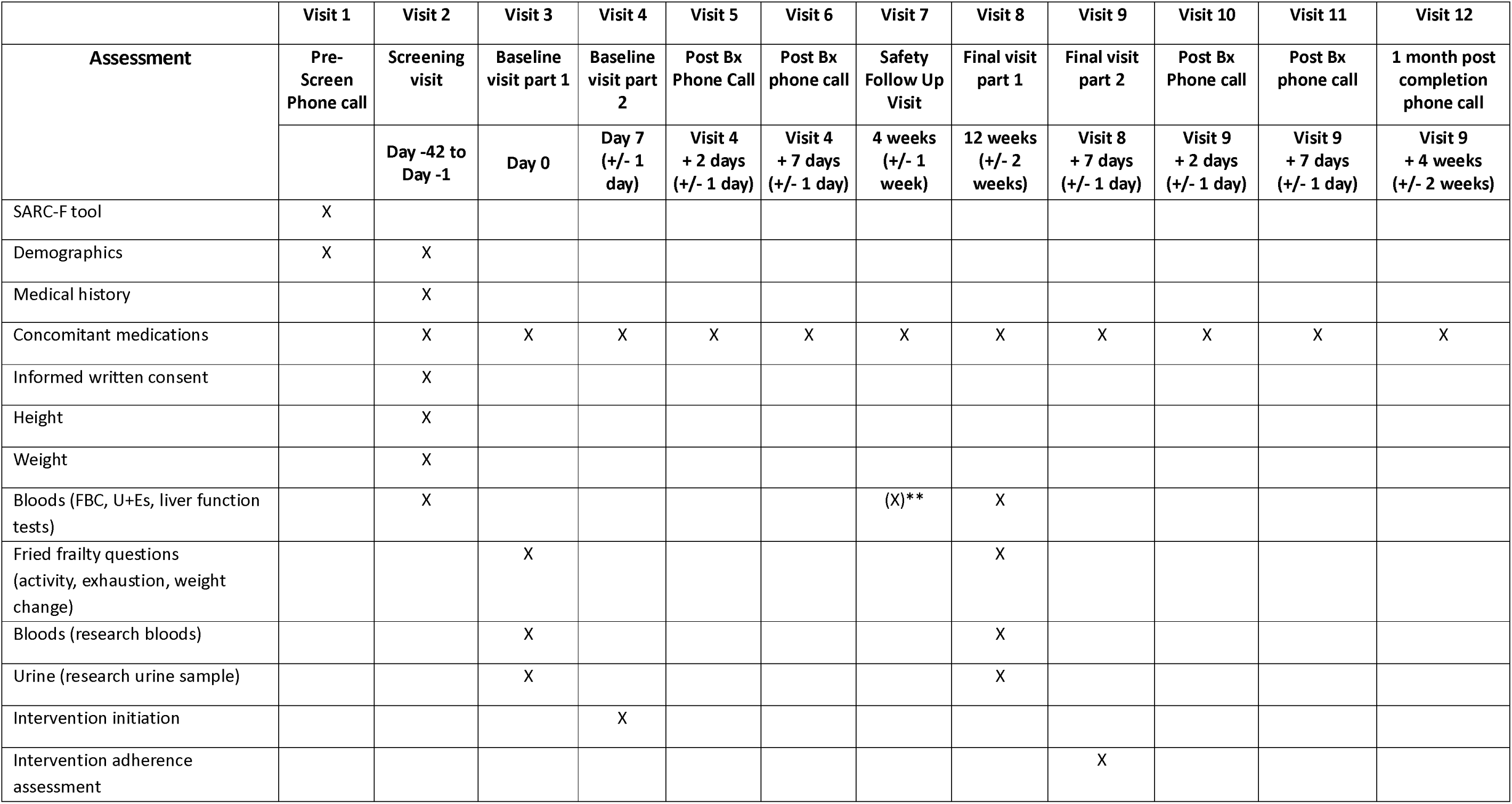
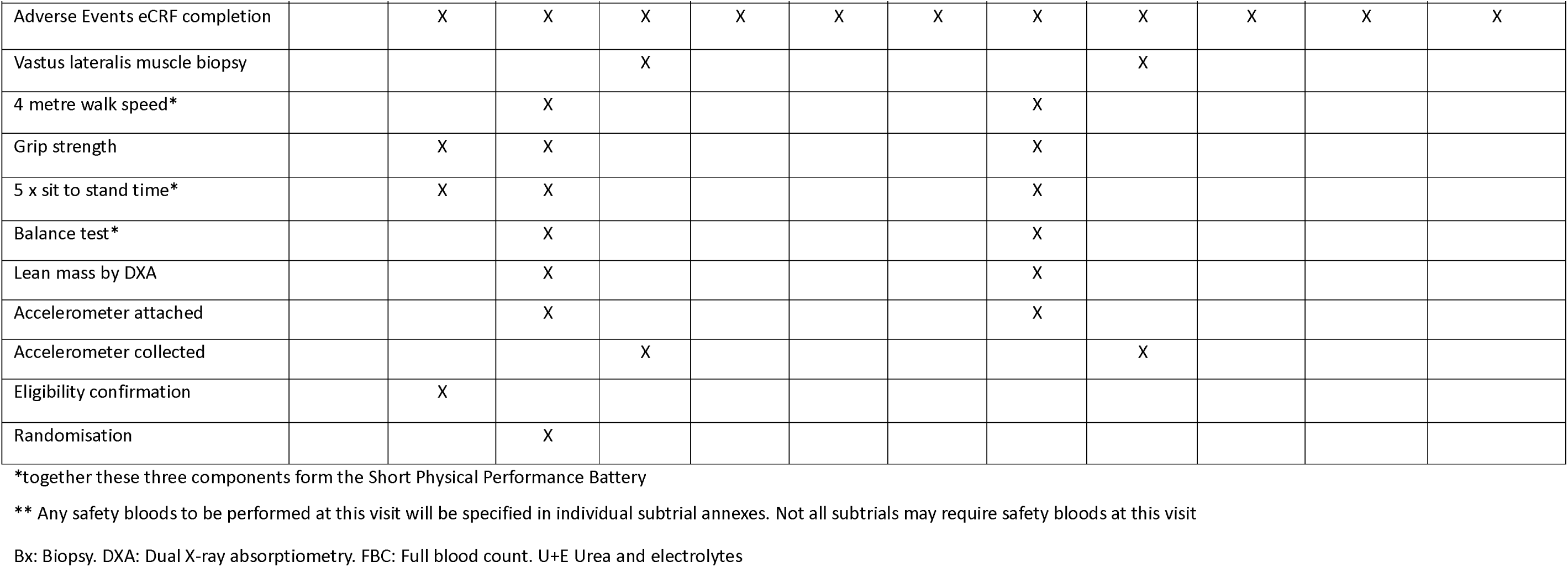
Schedule of Events.

### Sample size

The minimum clinically important difference for the 4 metre walk speed is 0.1m/s (25). Future large trials with heterogenous populations are likely to suffer from some dilution of effect size, and subtrials in this platform will be powered to detect a between-group difference in 4 metre walk speed at 12 weeks when the difference is 0.15m/s to ensure a robust proof-of-concept signal that justifies further testing in phase II trials. Assuming an standard deviation of 0.24 as seen in previous similar sarcopenia trials, and a correlation between baseline and 6 month measures of 0.84 as seen in a recent trial of metformin in older people with sarcopenia(20) (a similar population to those who will be enrolled in this trial), a total of 24 participants would be needed (12 per group) to give 80% power to detect this difference at alpha=0.05 (two-sided) in an analysis that adjusted for baseline values. To account for dropout, the default sample size for each subtrial will be 30 participants (15 per group). Different sample sizes may be stipulated for each subtrial in the subtrial annex, for example if a particular mechanistic outcome required a larger sample size, or if pooling of control groups across subtrials was planned with consequent efficiencies in required sample size.

### Recruitment

Potential participants will be identified through three main routes: research registries, secondary care, primary care. Registry sources include SarcNet (REC 18/NE/0314), the MULTIPLE registry (REC 22/NE/0014), and the NIHR BioResource Centre Newcastle (REC 18/NE/0138). Individuals listed in these registries have previously consented to be contacted about research opportunities and will be mailed an invitation letter and a brief study information sheet.

In secondary care, patients at Newcastle Hospitals and other participating sites acting as Participant Identification Centres (PICs) will be approached either face-to-face or by post, depending on the method of identification. In primary care, GP practices working with the NIHR Research Delivery Network will screen practice lists to identify eligible individuals and send study invitations and brief study information sheet by post.

Interested participants can respond by returning the reply slip in a pre-paid envelope or contacting the REVITALiSE research team directly by phone or email. All responses will be handled by the team based at the NIHR Newcastle Biomedical Research Centre. Individuals who express interest will be contacted for telephone pre-screening.

### Assignment of intervention and subtrial allocation

Where a single subtrial is open, the participant will automatically be allocated to that subtrial. Where more than one subtrial is currently open to recruitment, the investigator will ascertain which subtrials the participant is eligible to take part in. If the participant is eligible to take part in more than one open subtrial, the participant will be able to choose which subtrial to take part in after discussion with the investigator.

### Randomisation

Randomisation will be performed using a web-based randomisation system (REDCap). Randomisation within each subtrial will take place independently. Restricted randomisation using random permuted blocks will ensure balance across the two arms but randomisation will not be stratified. Randomisation will be performed after the first baseline visit assessments are complete. The default randomisation ratio will be 1:1 between the intervention and control arms unless otherwise specified in the subtrial annex information; when new subtrials are added, a decision will be made as to whether to pool control groups across subtrials, and if this takes place, there will be the opportunity to vary the control: intervention allocation ratio to optimise sample size whilst retaining power for the primary outcome.

### Data collection and management

All trial data will be collected via REDCap, entered directly into electronic case report forms (eCRFs) by trained site staff during study visits. The REDCap system will include built-in validation rules and range checks to minimise data entry errors and ensure completeness and consistency of the dataset. User access will be role-based, with permissions restricted according to study responsibilities to maintain data confidentiality and integrity. An audit trail will be maintained to track all data modifications. Routine data monitoring and quality assurance checks will be performed to identify and resolve discrepancies. All data will be stored on secure servers compliant with institutional and regulatory requirements, including Good Clinical Practice (GCP) and UK data protection legislation. The participant trial record will be archived at site following the end of the trial.

### Statistical analysis

Full details of all statistical analyses for each subtrial will be pre-specified in a statistical analysis plan (SAP) developed by the trial statisticians in conjunction with the Trial Management Group. This will include evaluation of clinical and mechanistic outcomes data. The analysis will be carried out in the mITT population. This population consists of all randomised participants within a given subtrial who undergo both baseline and follow up assessment for a given outcome, analysed according to their randomisation arm regardless of whether they received the allocated intervention or discontinued the intervention.

1. Analysis of the Primary Outcome Measure The main primary objective is to estimate the mean difference in the 4 metre walk speed at 12 weeks adjusted for baseline values in adults aged 65 and over with probable sarcopenia treated with an intervention compared to usual care, regardless of deviations from the allocated intervention for any reason. A linear regression model will be fitted to the 4 metre walk speed at 12 weeks adjusted for baseline 4 metre walk speed. The estimated adjusted mean difference between intervention and usual care at 12 weeks will be extracted from the model and reported with a 95% confidence interval.
2. Analysis of Secondary Outcome Measures Continuous secondary endpoints will be analysed as described for the primary outcome. Binary secondary endpoints will be analysed with a logistic regression model. All models will be adjusted for the measure at baseline (if taken).

### Assessment of Adherence

Adherence will be assessed as detailed in the trial annex. For example, for nutritional supplements, adherence will be calculated by subtracting the quantity of supplement returned (e.g., number of capsules or weight of supplement powder) from the quantity originally supplied, then dividing this by the quantity expected to have been consumed over the course of the trial. This adherence assessment will be conducted only at the end of the trial. Importantly, participants with low adherence will not be withdrawn from the study.

### Intervention discontinuation

An intervention will be discontinued in the following circumstances:

- Side effects that are possibly, probably or definitely related to the intervention and which are not tolerable to the participant
- A serious adverse reaction (SAR) or Unexpected Related Serious Event (URSE) occurs in a participant
- The participant requests the intervention to be discontinued.

The trial intervention may be temporarily discontinued at the instigation of the treating clinical team or the study team if necessitated by intercurrent illness.

Discontinuation of the intervention will not automatically lead to withdrawal from the subtrial, and participants who discontinue the intervention may still undergo the final study visit including muscle biopsy if the participant is willing to continue with trial follow-up.

### Withdrawal from trial

Participants will have the right to withdraw from the trial at any time without having to give a reason. Participants who withdraw from the trial once they have been randomised will not be replaced.

Where a participant withdraws from the trial, or is withdrawn by an Investigator, the data collected up to the point of withdrawal will be retained and included in the analysis. No additional research data will be collected from the participant after the point of withdrawal.

#### Participant Requested Withdrawal

When a participant states they want to withdraw from the trial, site staff should try to ascertain the reason for withdrawal and document this reason within the eCRF and participant’s medical notes. Although a withdrawal form is available for participants to complete, participants should not be pressurised to complete this. Participants may verbally indicate that they wish to withdraw from the trial.

#### Investigator Led Withdrawal

The Investigator may withdraw a participant from the trial at any time if they deem it necessary for the following reasons:

- Continuing to attend trial visits and undergoing trial assessments would place unreasonable demands on the physical or psychological health of the participant.
- The participant loses capacity for a prolonged period
- An adverse event, which results in inability to continue to adhere to trial procedures
- Unwillingness of the participant to engage with the trial visits or schedule
- Termination of the clinical trial by Sponsor

### Trial oversight

#### The Trial Management Group (TMG)

The Trial Management Group (TMG) will meet approximately once a month for the duration of the trial. Membership will include the Chief Investigator, co-applicants, trial statistician, local site research staff, and a representative from the Sponsor. The TMG will review all adverse events related to trial interventions or outcome measures as part of its regular agenda. In addition to its management functions, the TMG will also fulfil the role of the Trial Steering Committee. Given the low number of participants and the short duration of therapy, an independent Data Monitoring Committee will not be established. However, to ensure participant safety and data integrity, an interim review of study procedures, AE, SAE, treatment discontinuation, participant withdrawal rates and missing data, will be conducted by the Sponsor when 50% of participants have been enrolled into a given subtrial

### Patient and public involvement

A dedicated Patient and Public Involvement and Engagement (PPIE) advisory group was established at the outset of the trial and meets quarterly to provide ongoing input. The group has played a key role in advising on trial design, participant-facing materials, outcome measures—including the acceptability of biopsies—and the selection of trial interventions. Their involvement will continue throughout the duration of the platform, with consultation on future interventions and the development of intervention-specific outcome measures for each subtrial. The PPIE group is supported by the trial’s PPIE Manager, Chief Investigator, and Deputy Chief Investigator, who provide regular feedback to the Trial Management Group (TMG). This collaborative model of engagement was co-developed with the PPIE group themselves. The group will be actively involved in shaping strategies for trial dissemination.

### Harms

All adverse events (AEs) occurring from the point of consent until the end of trial participation will be documented. Falls themselves will not be considered AEs; however, any injuries resulting from a fall will be recorded as an adverse event. Mild pain or soreness within the first 48 hours after a muscle biopsy, as well as superficial bruising at the biopsy site, are expected and will not be classified as AEs. Investigations, interventions, procedures, and operations will not be recorded as AEs, but any new underlying conditions that necessitate these interventions will be documented as AEs. Additionally, any complications arising from investigations, interventions, procedures, or operations will be recorded as AEs.

### Consent

Written informed consent will be sought at the screening visit, prior to conducting any trial procedures, including screening assessments. Consent will be sought for participation in the main platform trial at the start of the screening visit, with additional consent obtained for a specific subtrial once it has been selected after the screening visit.

A member of the research team, competent in obtaining informed consent for research studies and formally delegated to do so by the site Principal Investigator, as evidenced by dated signatures on the site delegation log, will conduct a consent interview with the participant in the clinic or the participant’s home if preferred by the participant.

### Ancillary and post-trial care

Participants will continue to receive standard clinical care throughout their involvement in the trial, and any medical issues identified during the study will be managed in accordance with routine clinical practice. Where trial procedures uncover clinically relevant findings, participants will be referred to appropriate healthcare services for further assessment or treatment, as needed. Although no formal ancillary care beyond standard National Health Service (NHS) provision is planned, participants will have access to the research team for the duration of their involvement to address any trial-related concerns. Post-trial, participants will return to usual care pathways. Results of the trial will be shared with participants in an accessible format, and information on any future related studies will be provided where appropriate.

### Ethics and Dissemination

The trial is approved UK Health Research Authority and Northeast – Tyne and Wear South Research Ethics Committee (IRAS 352708; ref: 25/NE/0115) and is registered on the ISRCTN trials registry (ISRCTN10801475). Results will be made available to participants, their families, patients with sarcopenia, the public, regional and national clinical teams, and the international scientific community.

### Access to data and samples

Access to the full dataset will be limited to the Trial Management Group (TMG) and authors of the trial publication. At the conclusion of the trial, a de-identified dataset will be prepared and securely stored by Newcastle University. Requests for data sharing with external study teams, including international collaborators, will be considered on a case-by-case basis by a Data Access Committee comprising representatives from the Funder, Sponsor, and the trial Chief Investigator.

Participants are consented for the biobanking of biological samples, including optional consent for their future use in animal research and commercial studies. Requests to access stored samples after the completion of the trial will be reviewed by a Trial Access Committee in consultation with the Sarcopenia, Ageing and Multimorbidity Theme of the NIHR Newcastle Biomedical Research Centre, ensuring alignment with participant consent and research governance policies.

## Intervention selection process

A structured two-step process has been developed to identify and prioritise interventions for evaluation within the REVITALiSE platform and potential future platforms aimed at assessing pharmaceutical interventions. First, a comprehensive horizon scan has been conducted to identify promising pharmaceutical, nutraceutical, and technology-based interventions in early-stage development—defined as those not yet progressed to phase II or later clinical trials—targeting the prevention, delay, or treatment of sarcopenia. The inclusion criteria, databases searched, and search dates are summarised in Table 2, with the full search strategy detailed in the supplementary material. Records have been initially screened by a single researcher, with eligibility uncertainties resolved through team consensus. Ten percent of entries were independently verified for quality assurance.

**Table 2.** Horizon Scan search criteria.

|  |  |
| --- | --- |
| Intervention | Any pharmaceutical, nutraceutical and technology-based intervention to prevent, delay onset of, and/or treat sarcopenia.<br>Interventions that address muscle strength and/or muscle mass will be included.<br>Technologies may include but are not limited to electrical stimulation, sensor-based or wearables, vibration therapies. |
| Outcome | Any measure of sarcopenia, muscle mass (including muscle mass, lean mass or fat-free mass), muscle strength, or muscle quality.<br>Any mechanistic outcome pertinent to muscle strength and mass. |
| Study design | Preclinical (partial and full animal models, laboratory) studies, first-in-human studies, including studies that are ongoing or published in the last five years. |
| Databases and dates | Searches were performed using<br>Pubmed (searched from 1st January 2018 to 9th January 2024, English language filter applied);<br>MedRXiv (searched for preprints from 1st January 2018 to 12th January 2024, no language restrictions were applied);<br>BioRXiv (searched for preprints from 1st January 2018 to 12th January 2024, no language restrictions were applied);<br>Clinicaltrials.gov (searched 16th January 2024, with clinical trials filtered for trial phase 1, 1/2 and not applicable (N/A);<br>UK research funding sources, including UK Research and Innovation (UKRI); Medical Research Council (MRC); National Institute for Health and Care Research (NIHR); Wellcome Trust (searched from 1st January 2018 to 12th January 2024, no language restrictions were applied). |

A total of 3,835 records were screened, of which, 235 have met eligibility criteria (138 preclinical studies, and 97 clinical trials). Sample sizes in clinical trials ranged from 6 to 630 participants. Patients with sarcopenia represented the largest studied population (51 trials). Interventions evaluated included nutraceuticals (47%), pharmaceuticals (39%), and technology-based therapies (6%).

To prioritise candidates for evaluation within the REVITALiSE platform, a multidisciplinary team with expertise in sarcopenia, early-phase clinical research, and geriatric medicine (CM, RP, MW) developed a structured intervention selection algorithm. This tool combines “stop/go” criteria with traffic-light ratings to facilitate objective decision-making. The algorithm was applied to horizon scan data and refined iteratively based on the outcomes. The algorithm is presented in Figure 1 and Table 3.

**Table 3.**
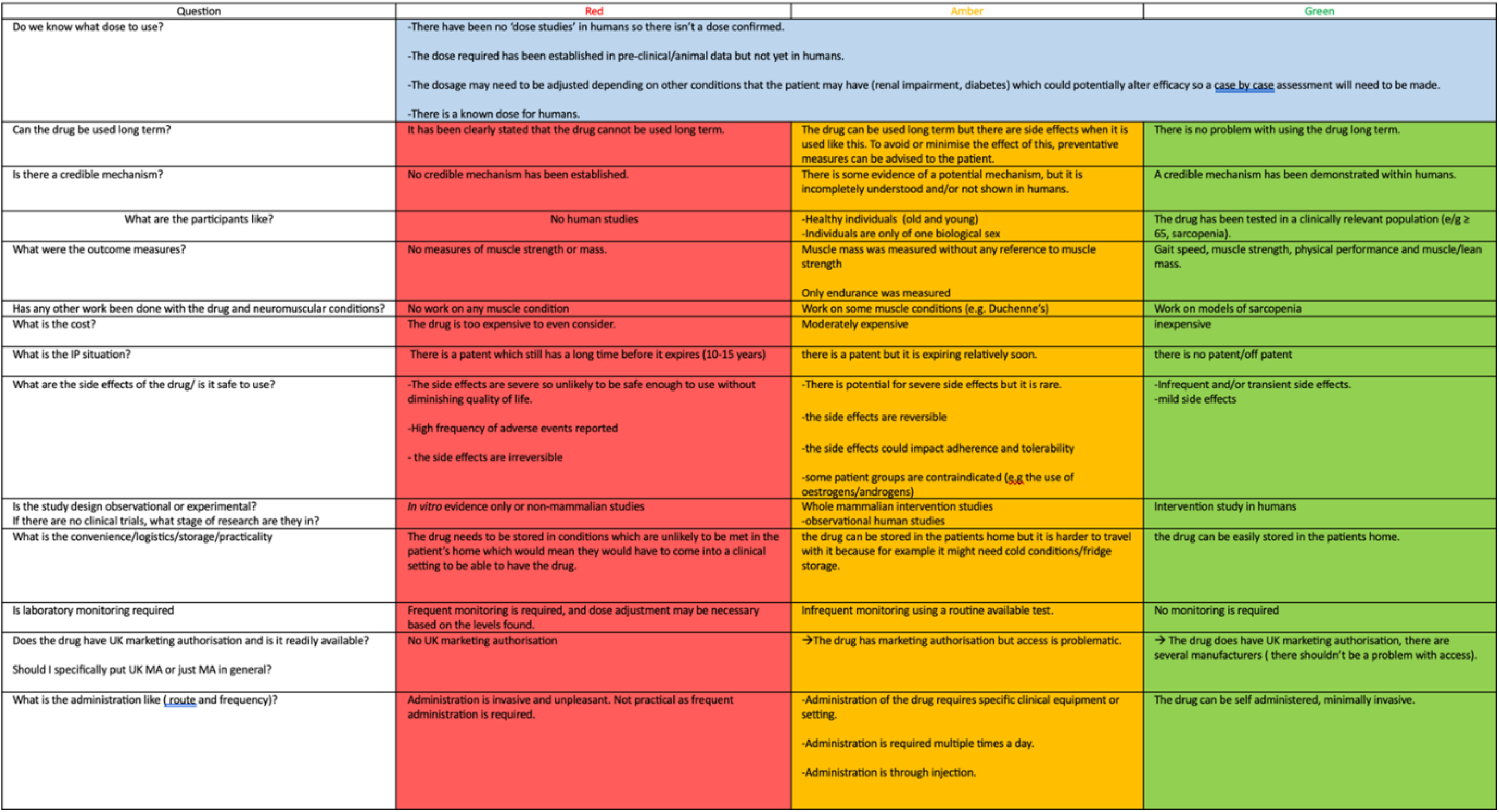
Red/Amber/Green (RAG) rating system for proposed interventions.

| Question | Red | Amber | Green |
| --- | --- | --- | --- |
| Do we know what dose to use? | -There have been no 'dose studies' in humans so there isn't a dose confirmed.<br>-The dose required has been established in pre-clinical/animal data but not yet in humans.<br>-The dosage may need to be adjusted depending on other conditions that the patient may have (renal impairment, diabetes) which could potentially alter efficacy so a <u>case by case</u> assessment will need to be made.<br>-There is a known dose for humans. |  |  |
| Can the drug be used long term? | It has been clearly stated that the drug cannot be used long term. | The drug can be used long term but there are side effects when it is used like this. To avoid or minimise the effect of this, preventative measures can be advised to the patient. | There is no problem with using the drug long term. |
| Is there a credible mechanism? | No credible mechanism has been established. | There is some evidence of a potential mechanism, but it is incompletely understood and/or not shown in humans. | A credible mechanism has been demonstrated within humans. |
| What are the participants like? | No human studies | -Healthy individuals (old and young)<br>-Individuals are only of one biological sex | The drug has been tested in a clinically relevant population (e/g ≥ 65, sarcopenia). |
| What were the outcome measures? | No measures of muscle strength or mass. | Muscle mass was measured without any reference to muscle strength<br>Only endurance was measured | Gait speed, muscle strength, physical performance and muscle/lean mass. |
| Has any other work been done with the drug and neuromuscular conditions? | No work on any muscle condition | Work on some muscle conditions (e.g. Duchenne's) | Work on models of sarcopenia |
| What is the cost? | The drug is too expensive to even consider. | Moderately expensive | Inexpensive |
| What is the IP situation? | There is a patent which still has a long time before it expires (10-15 years) | there is a patent but it is expiring relatively soon. | there is no patent/off patent |
| What are the side effects of the drug/ is it safe to use? | -The side effects are severe so unlikely to be safe enough to use without diminishing quality of life.<br>-High frequency of adverse events reported<br>- the side effects are irreversible | -There is potential for severe side effects but it is rare.<br>-the side effects are reversible<br>-the side effects could impact adherence and tolerability<br>-some patient groups are contraindicated (e.g the use of oestrogens/androgens) | -Infrequent and/or transient side effects.<br>-mild side effects |
| Is the study design observational or experimental?<br>If there are no clinical trials, what stage of research are they in? | *In vitro* evidence only or non-mammalian studies | Whole mammalian intervention studies<br>-observational human studies | Intervention study in humans |
| What is the convenience/logistics/storage/practicality | The drug needs to be stored in conditions which are unlikely to be met in the patient's home which would mean they would have to come into a clinical setting to be able to have the drug. | the drug can be stored in the patients home but it is harder to travel with it because for example it might need cold conditions/fridge storage. | the drug can be easily stored in the patients home. |
| Is laboratory monitoring required | Frequent monitoring is required, and dose adjustment may be necessary based on the levels found. | Infrequent monitoring using a routine available test. | No monitoring is required |
| Does the drug have UK marketing authorisation and is it readily available?<br>Should I specifically put UK MA or just MA in general? | No UK marketing authorisation | →The drug has marketing authorisation but access is problematic. | → The drug does have UK marketing authorisation, there are several manufacturers ( there shouldn't be a problem with access). |
| What is the administration like ( <u>route</u> and frequency)? | Administration is invasive and unpleasant. Not practical as frequent administration is required. | -Administration of the drug requires specific clinical equipment or setting.<br>-Administration is required multiple times a day.<br>-Administration is through injection. | The drug can be self administered, minimally invasive. |

## Discussion

Although our understanding of the biological mechanisms underlying skeletal muscle ageing has grown substantially, research activity in experimental medicine and early-phase clinical trials for sarcopenia remains limited (3). Establishing trial infrastructure that supports the early evaluation of novel interventions is crucial to translating these scientific discoveries into clinical practice. This platform trial is the first of its kind in sarcopenia and is designed to provide a practical model for future studies, as well as a framework to improve the conduct of early-phase sarcopenia research. In doing so, it aims to accelerate the development of new interventions and create a structured pathway into later-phase efficacy trials.

The platform’s flexible and innovative design distinguishes it from more traditional platform trials that often rely on a single, shared control arm (26). Instead, this trial adopts a more adaptable structure that maintains the methodological efficiencies and resource-sharing benefits of the platform model while allowing for tailored subtrial designs. This flexibility enables the evaluation of a diverse range of intervention types—including exercise-based, device-driven, and nutraceutical approaches—better aligning with the multifactorial nature of sarcopenia and the evolving landscape of potential therapies.

A key strength of the platform is the integration of mechanistic outcomes within each subtrial, which will significantly advance understanding of sarcopenia pathophysiology. By prioritising both mechanistic and clinical endpoints, the trial not only generates early signals of efficacy but also deepens insight into how specific interventions exert their effects. This dual focus enhances scientific knowledge and provides critical early data that can inform the design of later-phase studies, while also helping to de-risk future research investment from academic and commercial partners.

The trial builds on successful processes and outcome measures validated in previous studies such as MET-PREVENT, LACE, and the Acipimox trial, ensuring that the methodological foundation is robust (20, 21, 27). Furthermore, the platform incorporates an inclusive approach to participant engagement, informed by prior work in MET-PREVENT (7). A distinctive strength of this trial is the extensive involvement of patients and the public from the outset. The PPIE group has contributed meaningfully to trial design, documentation, and decision-making structures, and will continue to guide ongoing operations and dissemination strategies—an approach that remains rare in early-phase research. Importantly, the PPIE group also co-developed the framework for their own engagement within the platform, determining how they are consulted and involved across different trial activities. This collaborative model ensures that involvement is structured, consistent, and meaningful, setting a precedent for PPIE in early-phase and experimental medicine trials.

Another key feature is the structured and adaptable process developed to identify, evaluate, and prioritise candidate interventions for inclusion on the platform. This mechanism not only supports early-phase selection but can also be applied to inform the transition of promising interventions into larger, later-phase trials. While the current platform does not include drug interventions due to cost and logistical constraints, expanding to include pharmacological agents remains an ambition for future iterations. Similarly, adaptive trial elements were not incorporated due to the small sample sizes and short follow-up periods, but this remains a potential avenue for future development as the platform matures.

The first intervention selected for evaluation within the platform is fisetin, a senolytic compound with preclinical promise in age-related conditions, including muscle ageing (28). We anticipate recruitment for this subtrial to commence in late 2025, marking the beginning of a novel, scalable approach to experimental medicine in sarcopenia.

## Data Availability

No data currently generated. Statement of data availability will be included in publications of trial results

## Acknowledgements

CMcD, MDW PW, HA, KK and AAS, acknowledge support from the National Institute for Health and Care Research (NIHR) Newcastle Biomedical Research Centre (BRC). MDW and KK acknowledge support from the NIHR Newcastle Clinical Research Facility. The NIHR Newcastle BRC is a partnership between Newcastle Hospitals NHS Foundation Trust, Cumbria Northumberland Tyne and Wear NHS Foundation Trust, and Newcastle University. The views expressed are those of the authors and not necessarily those of the NIHR or the Department of Health and Social Care.

## Contributorship

Overall study conception and design: MDW, AAS, CM, PW

Statistical analysis design: AM, NW, JW

Co-production of public and patient engagement approach: HA, CM, MDW

Development of intervention selection tool: MP, CM, MDW

Horizon scan: GFS, SK, JN, EGR, HO’K, MDW

Recruitment and delivery design: PW, KK, CM, MDW

Manuscript preparation: CM, PW, MDW

Critical revision of manuscript: All authors

